# Limited Echocardiogram Acquisition by Clinicians Aided with Deep Learning: A Randomized Controlled Trial

**DOI:** 10.1101/2025.04.18.25326085

**Authors:** Andre Kumar, Evan Baum, Caitlin Parmer, John Kugler

## Abstract

**Background:** Deep learning (DL) programs can aid in the acquisition of echocardiograms by medical professionals not previously trained in sonography, potentially addressing access issues in underserved communities. This study evaluates whether DL-enabled devices improve limited echocardiogram acquisition by novice clinicians not trained on sonography.

**Methods:** In this single-center randomized controlled trial (2023-2024), internal medicine residents (N=38) without sonography training received a personal ultrasound device with (N=19) or without (N=19) DL capability for two weeks while caring for patients on a hospital ward. Participants were allowed to use the devices at their discretion for patient-related care. The DL software provided real-time guidance for probe placement and image quality assessment. The primary outcome was time to acquire a five-view limited echocardiogram. Measurements occurred at randomization and after two weeks, with all scans performed on the same standardized patient. Secondary outcomes included image quality using the modified Rapid Assessment for Competency in Echocardiography (RACE) scale and participant attitudes.

**Results:** At baseline, both groups had comparable scan times and image quality scores. At follow-up, the DL group demonstrated significantly faster total scan times (152 seconds [IQR 115-195] vs. 266 seconds [IQR 206-324]; p<0.001; Cohen’s D 1.7) and better image quality with higher RACE scores (15 [IQR 10-18] vs. 11 [IQR 7-13.5]; p=0.034; Cohen’s D 0.84). Trust in the AI features did not differ between the groups post-intervention.

**Conclusions:** Ultrasound machines with DL features may improve image acquisition times and image quality by novices not trained in sonography. These findings suggest DL algorithms could help address critical gaps in image acquisition by healthcare professionals.

## Introduction

There is a critical shortage of sonographers worldwide, resulting in delays for crucial ultrasound examinations across a variety of clinical specialties.^1–3^ In the United States, the demand for ultrasound imaging has increased by 55.1% while the educational capacity has only grown by 23.0% between 2011-2021.^3^ This imbalance necessitates innovative approaches to improve patient access to ultrasound testing. Potential solutions include machine-assisted acquisition technologies, training of ancillary medical staff to acquire basic images for remote interpretation, and artificial intelligence-augmented models to aid non-sonographers.^1,4,5^ Clinicians can also address this gap through the expanded use of point-of-care ultrasound (POCUS) to make expedient decisions on patient triage, risk stratification, or the need for more extensive diagnostic testing.^6–8^

Deep learning (DL) algorithms have demonstrated remarkable capability at both aiding in image acquisition and interpretation of ultrasound images.^1,5,9,10^ These algorithms can be deployed on ultra-portable devices connected to smartphones or tablets, enabling diagnostic assessments in settings with traditional access barriers.^6,7^ DL may aid ancillary medical staff in acquiring ultrasound images that can be interpreted by a radiologist or provide real-time diagnostic augmentation for clinical providers at the patient’s bedside.^11^

Previous investigations have shown that DL-assisted ultrasounds can help novices obtain high-quality cardiac images, accurately assess ventricular function, and identify pericardial effusions.^12,13^ Other applications include DL-augmented POCUS for diagnosing pediatric pneumonia^14^, deep venous thrombosis^15^, and pneumothorax.^16^ Despite evidence that non-sonographers can acquire diagnostic-quality images with DL assistance^12,13^, randomized studies comparing educational outcomes and the development of competency remain limited.^5^ Such investigations are crucial when considering widespread implementation of DL software in healthcare, particularly in remote or underserved areas.

We previously reported that ultrasound novices with DL-augmented devices could obtain a single cardiac view (apical-4-chamber) more efficiently without compromising image quality.^5^ This study assesses whether DL-augmented portable ultrasound devices improve novice scanning proficiency for a limited echocardiogram with multiple views. We utilized a different DL-augmented machine for this study to demonstrate the impact of DL on image acquisition with novices. We hypothesized that POCUS novices randomized to DL-enabled devices would demonstrate greater efficiency and proficiency than those without DL assistance.

## Methods

### Study Participants & Setting

We conducted a randomized controlled trial at a single academic institution from 09/2023-09/2024. Participants who were eligible were recruited via email. Our inclusion criteria included internal medicine residents rotating on the general inpatient wards service. We excluded residents who had taken an ultrasound elective offered by our residency program. At the time of the study, there was no formal POCUS credentialing pathway within the residency program. The Stanford University Institutional Review Board approved this investigation. This study was registered with ClinicalTrials.gov (Identifier: NCT05900440).

### Study Design

Recruited residents (N=38) were randomized 1:1 to receive a POCUS device with DL-functionality (Lumify™ + Ultrasight™, N=19) or without (Lumify™ alone, N=19) for two weeks (Figure 1). Outcomes were assessed at baseline and at two-week follow-up. Participants could use the devices at their discretion for patient care or self-directed learning.^5,17^ At the time of randomization, all participants received written and verbal instructions on their device’s functionality and access to online learning modules regarding POCUS acquisition and interpretation.^5^ Participants were instructed to save an image each time they used the device to track its frequency of usage. For privacy reasons, patient images were not reviewed by researchers, and no feedback was provided on participants’ scans.

**Figure 1.**
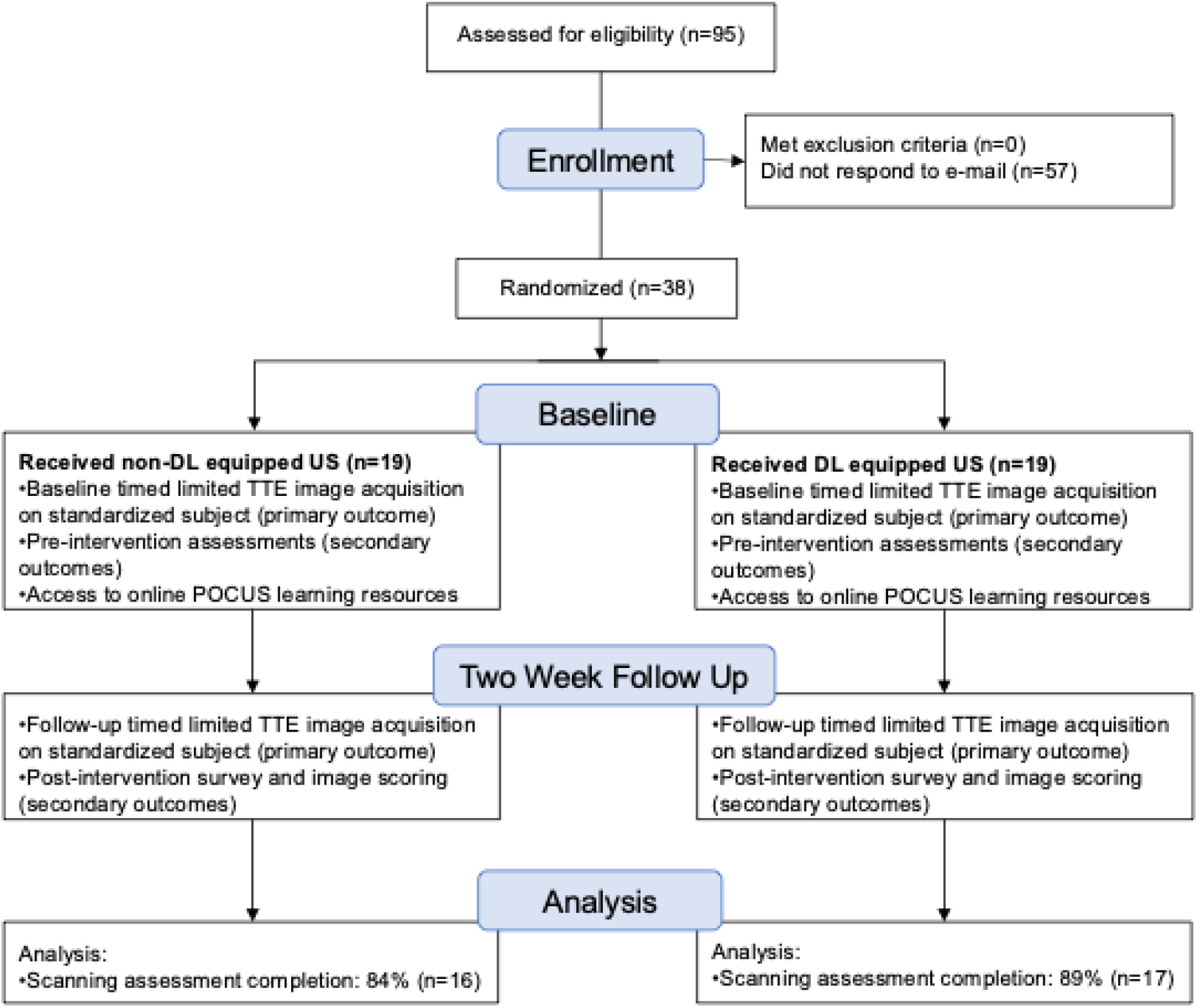
Study Design. TTE, transthoracic echocardiogram; DL, deep learning.

### Devices and Deep Learning Software

This study utilized the Philips Lumify™ portable ultrasound device. Participants in the DL group used the Ultrasight™ software, which interfaces with the Lumify display to provide real-time guidance for optimal probe placement to acquire basic cardiac views: parasternal long axis (PLAX), parasternal short axis (PSAX), apical-4-chamber (A4C), subcostal (SC), and inferior vena cava (IVC) views (Figure 2). The non-DL device did not provide these features. Additional information about the DL software can be found in the Supplementary Material.

**Figure 2.**
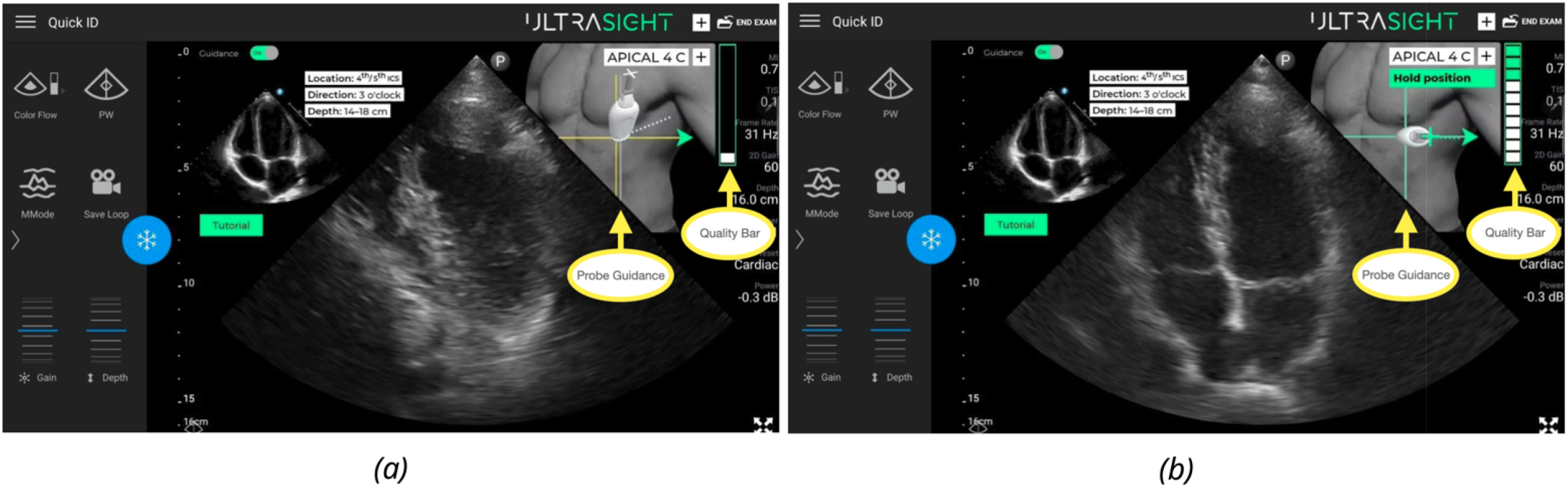
Deep Learning (DL) Software Overlay. The system uses AI guidance that overlays over the standard Philips Lumify display to aid the user in image acquisition (yellow arrows).. The figure shows two possible states of the system: navigation for an optimal view (Panel A), and holding the position once an optimal view has been obtained (Panel B). The display is continuously updated while the user performs the exam.

### Outcomes

Our primary outcome was the total time to acquire a limited echocardiogram based on the five core views at two-week follow-up: PLAX, PSAX, A4C, SC, and IVC. Participants were not assessed on doppler imaging studies as part of their acquisition. Secondary outcomes included the quality of the images acquired at follow-up and participant attitudes.

### Assessments/Surveys

Measurements were performed at the time of randomization (baseline) and at two-week follow-up. We performed assessments utilizing a different ultrasound device (Butterfly IQ+) to assess if the acquired skills used during the intervention period would transfer to other devices, which could represent a true improvement in performance rather than familiarity with a single device. All of the scanning assessments were performed on the same standardized patient with the same device. A study author was present for the scanning assessments to provide instruction and to set up the device, but they did not directly observe or comment on the images being acquired.

For the primary outcome of scanning time, participants were instructed to notify the proctor when they had acquired each view. Participants were timed from the moment the probe touched the patient’s torso until proctor notification that the exam was completed. The time to acquire each view was tracked during the assessment. Participants were instructed to save a five second video clip for each of the views for image quality analysis.^5,17^ Participants were instructed to notify a proctor if they decided to skip or give-up on acquiring a view, which was tracked. Participants could spend unlimited time attempting to acquire a particular view.

For image quality assessments, we used the modified Rapid Assessment of Competency in Echocardiography (RACE) scale, which rates each view on a 1-5 ordinal scale (Supplementary Material). The RACE tool has excellent interrater reliability (α = 0.87) and has been previously validated as an assessment tool for image acquisition and quality with POCUS.^18–20^ Three reviewers (AK, EB and JK), who were blinded to the study arms, independently reviewed the baseline and follow-up assessment scans to assign RACE scores. The average scores between the three reviewers were used to create composite RACE scores for analyses.

Surveys were administered to the participants at follow-up. These surveys assessed trainee attitudes toward POCUS, trust in the AI system, and their own confidence in acquiring images. Attitudes were measured using 5-point Likert scales. These surveys were based on previously described assessments.^17,21^

### Statistical Analysis

An a priori power analysis determined that 38 participants (19 per group) would provide >80% power to detect a 30% difference in the primary outcome between groups (Cohen’s d=0.73, two-sided alpha=0.05). The mean and SD assumptions were based on unpublished data collected at our center as part of two previous investigations, a recent investigation utilizing an A4C view and artificial intelligence (AI), as well as our experience in teaching cardiac POCUS to novices.^5,17,21^ Within the limitations of these assumptions, this study was sufficiently powered to detect a difference in the primary outcome.

We compared baseline performance and attitudes between groups. Scan time distributions were visualized with box/violin plots. We reported median and interquartile range (IQR) by randomized group and performed Mann-Whitney U testing to compare medians at follow-up. Similar analyses were conducted for all secondary outcomes. Cohen’s D measured effect size for significant differences, and Chi-square tests compared confidence levels.

All statistical analyses were performed using Python (Python Software Foundation, 2024), with packages including NumPy, pandas, and SciPy. ^22–24^

## Results

### Baseline Characteristics

There were a total of N=95 residents eligible for participation, of which N=38 responded to the email invitation to participate. No residents were excluded. N=19 were randomized to the DL-enabled device, and N=19 were randomized to the non-DL device. There were N=33 residents (87%) who completed the two-week follow up assessment, which included N=17 (89%) DL residents and N=16 (84%) non-DL residents (Table 1). Participant demographics and their prior POCUS experience are shown in Table 1.

**Table 1.**
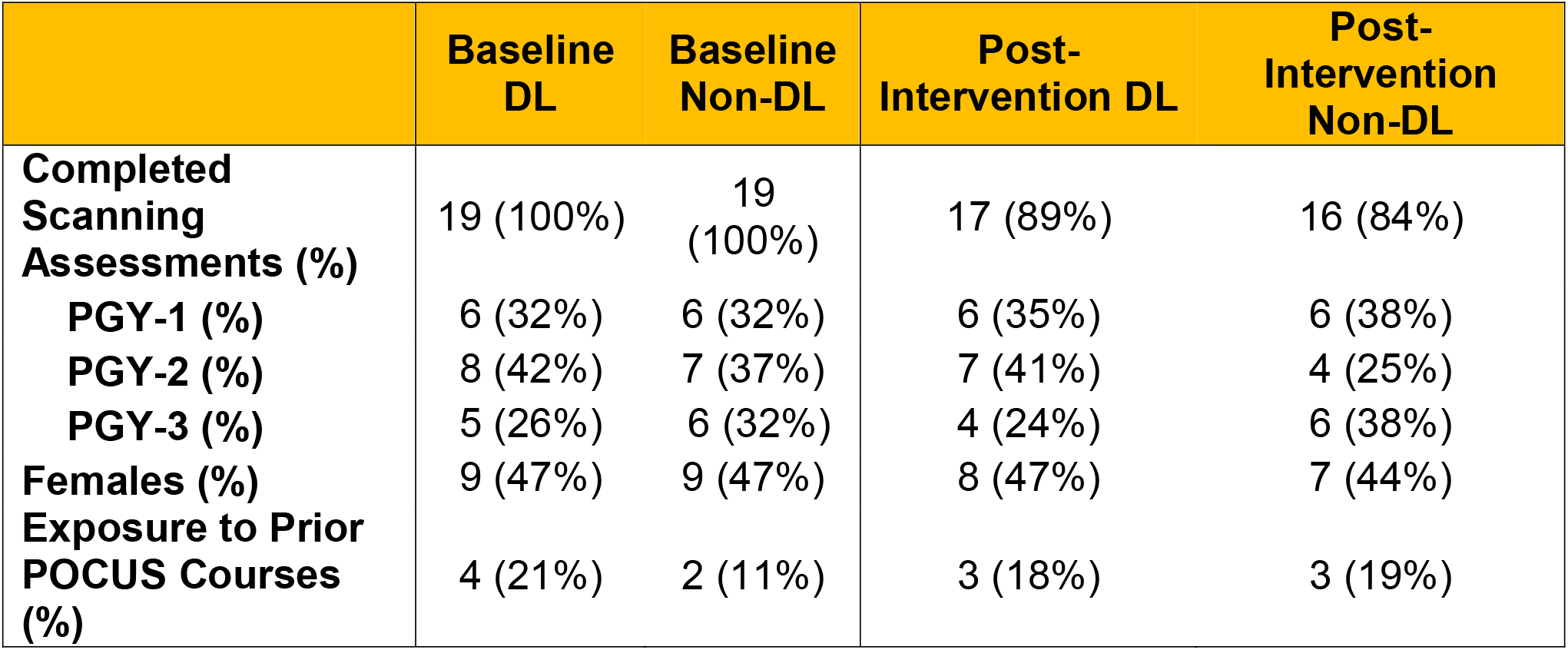
Participant Demographics. PGY, post-graduate year; POCUS, point-of-care ultrasound

### Device Usage

Participants in both arms were tracked on the frequency they used the devices using an electronic log (see Methods). Participants in both arms reported similar device usage rates (DL mean 5.4 times [SD 3.2] vs. non-DL mean 4.8 times [SD 2.9]; p=0.59).

### Primary Outcome: Time to Scan

Both groups had statistically similar baseline times to acquire limited echocardiograms (DL median 239s [IQR 146-267] vs. non-DL median 264s [IQR 151-332]; p=0.31). At follow-up, the DL group demonstrated significantly faster scanning times with a large effect size (median 151s [IQR: 115-195]) compared to the non-DL group (median 267s [IQR: 206-325]; p<0.001; Cohen’s D=1.7; Table 2).

**Table 2.** Study Outcomes. PLAX, parasternal long axis; PSAX, parasternal short axis; A4C, apical four chamber; SC, subcostal axis; IVC, inferior vena cava axis; DL, deep learning; IQR, interquartile range.

|  | Overall | DL Group | Non-DL Group | P-Value |
| --- | --- | --- | --- | --- |
| **Time to Acquire View, Median Seconds (IQR)** |  |  |  |  |
| Overall | 204 (152-291) | 151 (115-195) | 267 (206-325) | **<0.001** |
| PLAX | 44 (20-57) | 23 (14-34) | 56 (49-68) | **<0.001** |
| PSAX | 26 (16 -50) | 20 (14-39) | 37 (17-54) | 0.36 |
| A4C | 53 (38-86) | 39 (32-48) | 84 (58-97) | **<0.001** |
| SC | 33 (19-49) | 31 (20-46) | 37 (19-65) | 0.28 |
| IVC | 27 (19-41) | 26 (18-35) | 27 (19-43) | 0.77 |
| **Modified RACE Score, Median (IQR)** |  |  |  |  |
| Overall | 12 (8-16) | 15 (10-18) | 11 (7-14) | **0.034** |
| PLAX | 3 (1-3) | 3 (2-3) | 2 (1-3) | 0.14 |
| PSAX | 2 (2-3) | 3 (2-4) | 2 (1-2) | **0.015** |
| A4C | 2 (1-3) | 3 (1-3) | 1 (1-2) | **0.038** |
| SC | 3 (1-3) | 3 (1-3) | 3 (1-3) | 0.55 |
| IVC | 3 (1-4) | 3 (1-4) | 3 (2-4) | 0.12 |
| **Device Usage Rates, Mean (SD)** | 5.1 (3.2) | 5.4 (3.2) | 4.8 (2.9) | 0.59 |

This difference in time was driven by faster scanning times for two views: the PLAX view (DL median 23s [IQR 14-34] vs. non-DL median 56s [IQR 49-68]; p<0.001, Cohen’s D=1.5) and the A4C view (DL median 39s [IQR 32-48] vs non-DL median 84s [IQR 57-97]; p<0.001; Cohen’s D=1.1). The other views (PSAX, SC, and IVC) showed no significant differences in scan times at follow-up (Table 2).

### Secondary Outcomes

#### A. Image Quality

Both groups had statistically similar median RACE scores at baseline (DL median 13 points [IQR 8.5-14.5] vs. non-DL group median 11 points [IQR: 8.5-15]; p=0.98). At follow-up, the DL group achieved significantly higher median RACE scores (15 points [IQR 10-18]) versus the non-DL group (11 points [IQR 7-13.5]; p=0.034; Cohen’s D=0.84; Table 2).

The differences in image quality were primarily from the PSAX view (DL median 3 points [IQR 2-4] vs. non-DL median 2 points [IQR 1-2]; p=0.014; Cohen’s D=0.99) and A4C view (DL median 3 points [IQR 1-3] vs. non-DL median 1 point [IQR 0.5-2]; p=0.038; Cohen’s D=0.87). All other views did not have statistically significant differences in RACE scores for image quality (Table 2).

#### B. Completed Scans

At baseline, both groups had statistically similar rates of incomplete views (DL: N=18 residents vs. non-DL: N=4 residents; Chi-Square 1.42, p=0.23). At follow-up, the DL group had significantly lower failure rates (DL: N=1 resident vs. non-DL: N=8 residents; Chi-Square 5.77; p=0.016). Failure rates across both groups on the follow up scans by view were: PLAX (3.2%), PSAX (3.2%), A4C (16%), SC (3.2%), and IVC (3.2%).

#### C. Survey Results

N=20 residents (53%) completed an optional survey at the end of the study to assess their attitudes toward DL and POCUS. Further sub-analysis by intervention arm was not performed. Of these respondents, 50% agreed or strongly agreed they would generally trust DL functionality on ultrasound devices. Additionally, 70% would trust real-time guidance for probe positioning using DL, and 70% would trust auto-interpretation features of DL on ultrasound machines.

## Discussion

As AI applications for medical imaging continue to expand, DL-enabled ultrasound devices can improve access to diagnostic testing for underserved areas by allowing non-sonographers to acquire crucial diagnostic information.^1,5,10,12,13,25,26^ This randomized study found that novice POCUS users with DL-enhanced devices obtained limited echocardiograms more proficiently and with higher quality images suitable for diagnostic interpretation compared to users with standard devices.

Our results expand on previous investigations in several domains. Previous investigations have shown that DL-assisted ultrasounds can aid in image acquisition and interpretation by non-sonographers^12,14^, but there have been few randomized trials.^5^ While we previously demonstrated improved performance for a single cardiac view with DL^5^, this study was conducted with a different device and DL-software and demonstrates that novices can achieve improved proficiency and image quality for a clinically-important examination (the limited echocardiogram). Unlike approaches using DL-enabled devices for blind sweeps^1^, our study demonstrates that novices can be directed to acquire images suitable for immediate interpretation, tele-monitoring, or analysis by DL algorithms. These findings support multiple approaches to address worldwide gaps in diagnostic medical imaging.^2,3^

Previous investigations with non-AI enabled ultrasound devices have found that it takes 20-30 scans to reach a pre-specified competency level for cardiac POCUS.^18,27–29^ In our study, users performed fewer scans (approximately 5 per group) and yet the DL-group was able to demonstrate superior performance in acquisition time and image quality. In contrast, we have previously demonstrated that giving novices ultrasound devices without AI does not improve acquisition or scanning quality.^17^ These results demonstrate that DL-enabled technologies can be used to enhance proficiency in performance and may serve as an alternative to traditional teaching methods that are difficult to scale or access due instructor shortages.^30–32^ While our results support the hypothesis that DL-enabled devices can improve POCUS learning among novices, it is important to consider whether such trainees would be considered “competent” with limited echocardiography compared to sonographers who must undergo thousands of hours of training and years of deliberate practice.^5,29,33,34^

In this study, our primary outcome of total scan time was primarily influenced by two views: parasternal long axis (PLAX) and apical-4-chamber (A4C). In our experience, learners have more difficulty in obtaining these views as they are more dependent on body habitus, positioning, and scan angle. In previous investigations that examined learning curves with cardiac POCUS among novices, the participants had lower image quality scores in the PLAX and A4C views compared to others, suggesting difficulty in obtaining the view.^17^ Similarly, we observed image quality scores (RACE) were mainly driven from the parasternal short axis (PSAX) and A4C views. Based on our experience in teaching cardiac POCUS to novices, these two views are more susceptible to poor scanning angles (resulting in off-axis views), which can quickly degrade image quality.^17^ There can also be an argument on whether the statistically significant differences in scan times and their effect sizes would result in clinically meaningful differences for patients or providers. We would contend that any improvement in scan time that does not compromise image quality supports the feasability of this technology to be widely implemented across non-traditional environments to improve patient access.

### Limitations

There are several limitations of this study. It was conducted at a single academic institution, which limits its generalizability. While our initial sample size calculation used parametric assumptions, the primary analysis employed non-parametric tests due to non-normal data distribution, which may have affected the statistical power to detect the hypothesized effect size. Participants were not blinded to their study arm, and the short follow-up period limits conclusions about long-term competency development. We did not provide feedback on image quality or acquisition, even though this is an effective means of teaching POCUS.^35^ Our scanning assessments were performed on a standardized patient with adequate cardiac windows. Therefore, these findings may not reflect real world practice wherein clinicians obtain images and evaluate them at bedside, often in patients with difficult anatomy. Nevertheless, these results represent an intriguing implementation of DL-enabled POCUS, with future studies being warranted to investigate its applications in medical training and cardiac image acquisition.

## Conclusions

POCUS novices randomized to DL-enabled devices for two weeks demonstrated faster image acquisition and higher image quality scores. As DL-enabled devices continue to improve, further studies should evaluate the level of expertise they can develop in novice scanners and their impact on patient care. Future research should focus on image acquisition in actual patients and whether novice-acquired images improve diagnostic access for underserved populations.

## Data Availability

The datasets generated and/or analyzed during the current study are not publicly available due to sharing limitations from our IRB protocol, but are available from the corresponding author on reasonable request.

## Acknowledgements

**None Word Count**

## Abbreviations

A4C: (Apical-4-chamber)
AI: (artificial intelligence)
POCUS: (point-of-care ultrasound)
RACE: (rapid assessment for competency in echocardiography)
DL: (deep learning)

