## Supplementary Material for "Limited Echocardiogram Acquisition by Clinicians Aided with Deep Learning: A Randomized Controlled Trial"

**Supplementary Figure 1: Detailed Description of Ultrasight System**

The main features of the UltraSight AI Guidance system (Quality Bar, View Detection, Probe Guidance) are implemented using a neural network. The user chooses a target view of the heart, from a list of the 10 supported standard cardiac views.

A user scans a subject using Philips Lumify US probe,. Based on the ultrasound images, the UltraSight AI Guidance guides the user on where to place the transducer and how to manipulate it to acquire an optimal view.

The guidance graphical instructions are continuously updated while the user moves the transducer. The system displays an image quality bar that is continuously updated while the user scans the patient, the user attempts to find the maximal quality.

When the user decides to acquire a clip, they use the Lumify application clip saving functionality to save a clip as in a standard echocardiography exam.

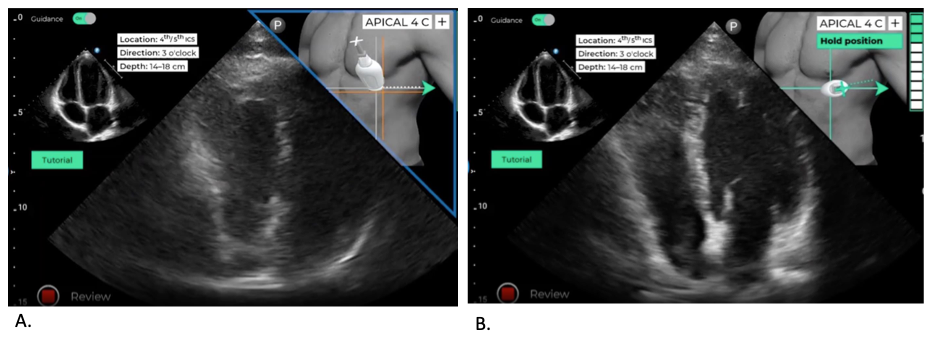

**Panel A** demonstrates that the user is off axis, and the system provides a suggestive corrective maneuver to obtain the optimal image. **Panel B** demonstrates that an optimal view has been obtained.

**Supplementary Figure 2. Modified Rapid Assessment for Competency in Echocardiography (RACE) Tool.**

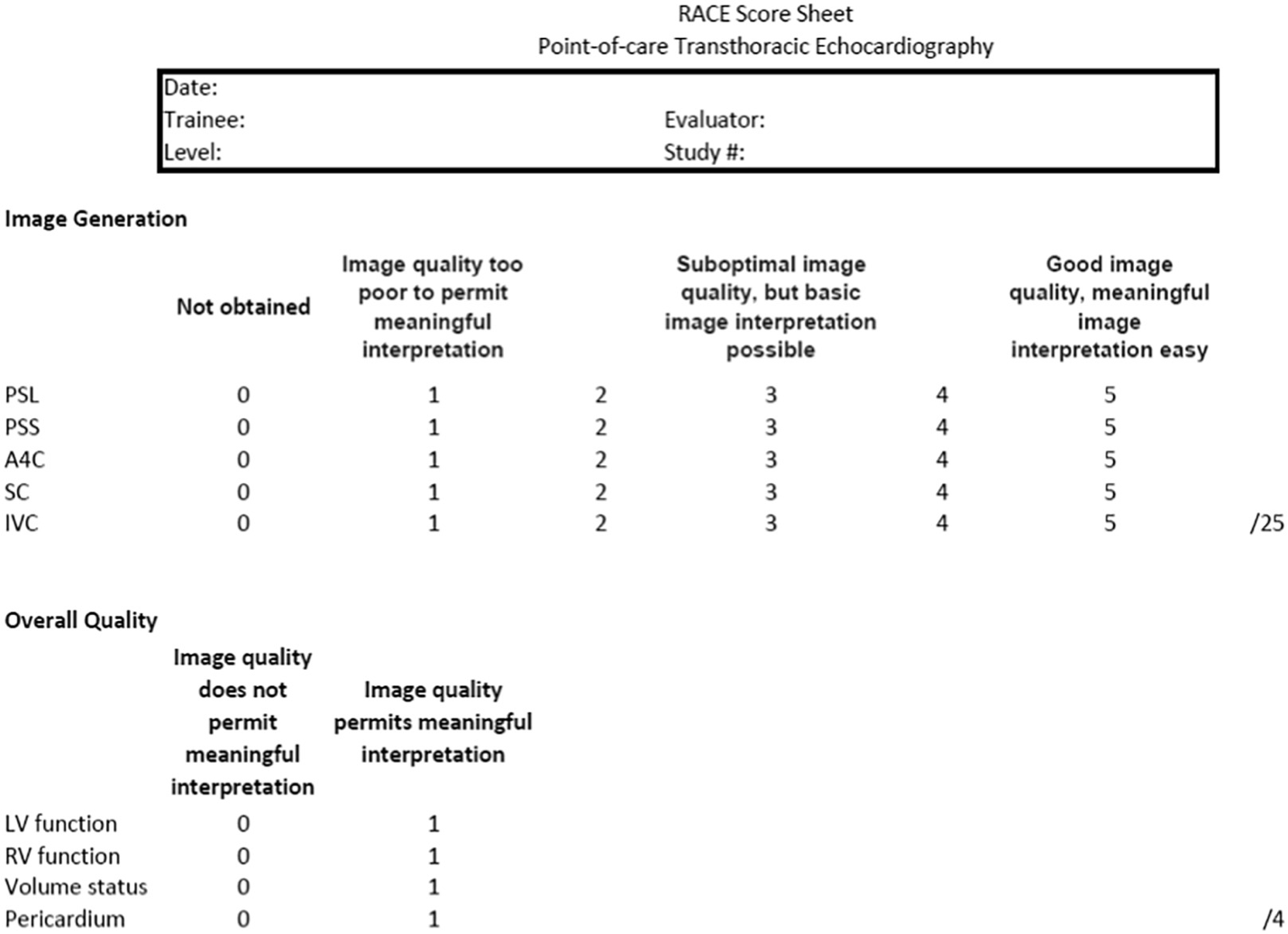
